# Longitudinal Associations Between Intrinsic Motivation and Subsequent Postpartum Health Behaviors and Life’s Essential 8: Results of the Pregnancy, Lifestyle and Environment Study-2 (PETALS-2) Cohort

**DOI:** 10.64898/2026.08.13.26360418

**Authors:** Brooke E. Wickman, Bridgette P. Smith, Michaela Kiernan, Monique M. Hedderson, Samantha F. Ehrlich, Charles P. Quesenberry, Andrea Millman, Hillary Serrato Bandera, Abigail Arons, Assiamira Ferrara, Susan D. Brown

**Affiliations:** Department of Internal Medicine, University of California, Davis, Sacramento, CA, USA; Stanford Prevention Research Center, Stanford University School of Medicine, Stanford, CA, USA; Division of Research, Kaiser Permanente Northern California, Pleasanton, CA, USA; Center for Upstream Prevention of Adiposity and Diabetes Mellitus (UPSTREAM), Division of Research, Kaiser Permanente Northern California, Pleasanton, CA, USA; Department of Kinesiology, Recreation, and Sport Studies, The University of Tennessee, Knoxville, Knoxville, TN, USA; Department of Pediatrics, University of California, Davis, Sacramento, CA, USA

**Keywords:** cardiovascular health, health behavior, intrinsic motivation, women’s health, postpartum

## Abstract

**Background:** Cardiovascular health is affected by health behaviors, but postpartum behavioral influences are not well understood. We examined whether intrinsic motivation (IM) is longitudinally associated with long-term postpartum health behaviors (healthy eating, physical activity, self-weighing) and cardiovascular health ((Life’s Essential 8 [LE8] scores).

**Methods:** The prospective Pregnancy, Lifestyle and Environment Study-2 (PETALS-2) followed women enrolled in the PETALS study at Kaiser Permanente Northern California during pregnancy (*N*=311). Data were collected via validated self-report surveys and objective measurements during pregnancy and 6-24 months postpartum (2017-2021). Health behaviors were dichotomized by sample-specific 75^th^ percentiles (*P75*) or pre-specified thresholds (attaining guideline-recommended moderate-to-vigorous physical activity [MVPA, ≥150 minutes/week]; self-weighing regularly [≥once/week]). Separate analyses lagged IM by timepoint to assess longitudinal associations between behavior-specific IM and immediate subsequent health behaviors; and between an IM composite and immediate subsequent LE8 scores.

**Results:** Each one-unit higher IM score was associated with greater likelihood of Healthy Eating Index-2015 scores ≥*P75* at 24 months postpartum (RR=1.42; 95% CI=1.07, 1.88); attaining MVPA guidelines at 6 (1.48; 1.03, 2.12), 12 (1.85; 1.26, 2.71), and 24 months postpartum (1.66; 1.22, 2.27); and regular self-weighing at 6 (1.53; 1.03, 2.27) and 12 months postpartum (1.65; 1.15, 2.36). Each one-unit higher composite IM score was associated with higher LE8 scores at 6, 18, and 24 months postpartum (18-month mean estimate=2.34; 95% CI=0.67, 4.02).

**Conclusions:** Greater IM was associated with healthier behaviors and cardiovascular health through 24 months postpartum. Future research should test whether interventions targeting IM improve health behaviors and long-term maternal cardiovascular health.

## Introduction

Healthy lifestyle behaviors that promote cardiovascular health include consuming a healthy diet, engaging in adequate physical activity, and achieving and maintaining a healthy weight. Diet, physical activity, and body mass index (BMI), among other health behaviors and factors, comprise the American Heart Association’s Life’s Essential 8 (LE8) score representing cardiovascular health.^1^

Women’s long-term cardiovascular health is influenced throughout life, and postpartum is a particularly susceptible yet opportunistic period.^2,3^ For example, weight retention after pregnancy increases the risk of later cardiovascular disease.^4,5^ However, randomized controlled trials testing behavioral interventions aimed at improving postpartum weight management via diet and physical activity have demonstrated only modest effectiveness.^6^

Despite the postpartum period’s influence on cardiovascular health,^3,7^ motivation for such behaviors has been understudied. Intrinsic motivation, defined as the extent to which individuals enjoy, value, and engage in behaviors with ease, is characterized as autonomous motivation driven by personal inclinations rather than external influence.^8–10^ Despite being modifiable,^11–13^ intrinsic motivation for healthy lifestyle behaviors has received limited research attention, particularly in postpartum populations.^14^ Prior work has identified a longitudinal association between greater intrinsic motivation and lower postpartum weight,^15^ in addition to cross-sectional associations between intrinsic motivation and health behaviors in pregnancy.^16^ However, research has yet to indicate whether intrinsic motivation is independently and prospectively associated with clinically meaningful engagement in cardiovascular health behaviors, or with quantifiable assessments of cardiovascular health through the extended postpartum period. Such findings could inform whether future behavioral interventions should focus on intrinsic motivation to potentially improve maternal cardiometabolic health.

To address current evidence gaps, the prospective Pregnancy, Lifestyle and Environment Study-2 (PETALS-2) aimed to identify motivational determinants of health behaviors from pregnancy through postpartum. We examined whether repeated measures of intrinsic motivation for healthy eating, physical activity, and self-weighing, each assessed separately, were longitudinally associated with subsequent engagement in corresponding behaviors throughout the postpartum period. We also examined whether intrinsic motivation was longitudinally associated with subsequent LE8 scores, as a comprehensive assessment of cardiovascular health (Figure 1).

**Figure 1.**
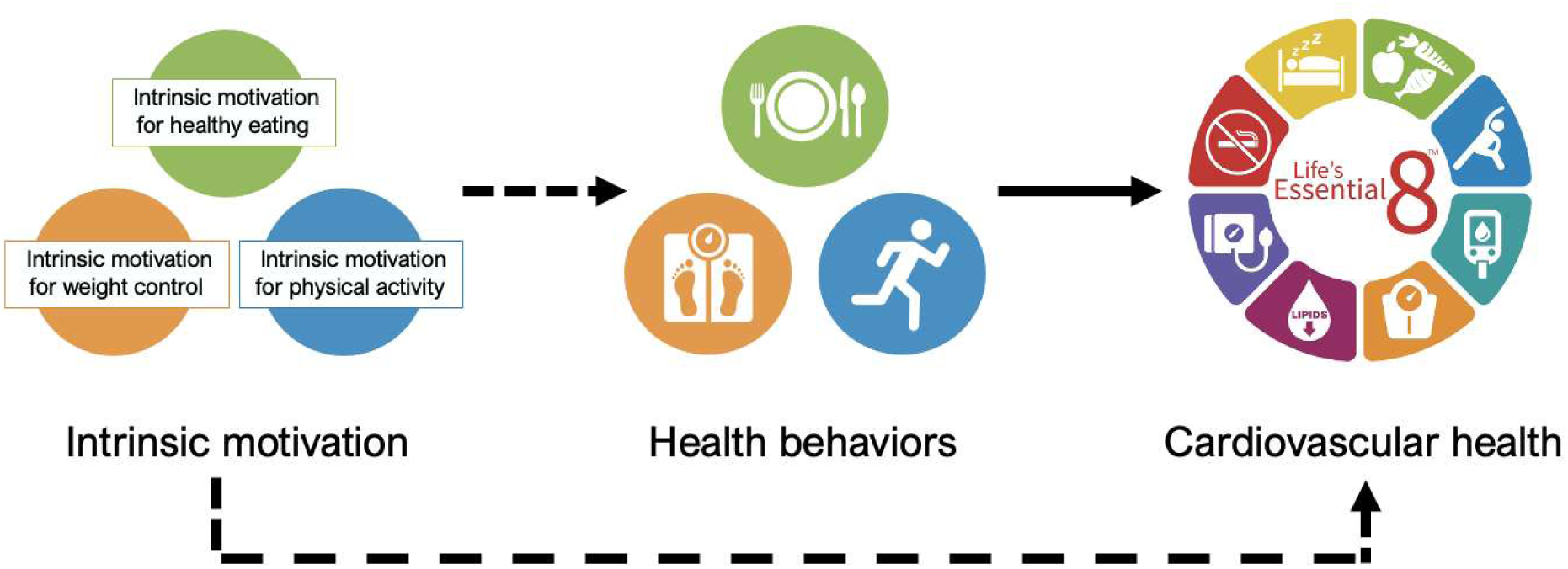
Conceptual model of the potential longitudinal association between intrinsic motivation, health behaviors, and life’s Essential 8 cardiovascular health scores Cardiovascular health is represented here as American Heart Association Life’s Essential 8 scores (pending copyright permissions from the American Heart Association).^1^

## Methods

### Study sample and procedure

PETALS-2 methods have been previously described.^15^ Briefly, participants were first enrolled in PETALS,^17^ a population-based study among healthy individuals aged 18-45 years during a singleton pregnancy in the Kaiser Permanente Northern California (KPNC) integrated healthcare system. In 2017-2018, questionnaires on IM and self-efficacy were added to those related to self-report demographics, clinical characteristics, and health behaviors that were collected during early pregnancy (gestational age 10-13 weeks). PETALS participants were potentially eligible for PETALS-2 postpartum recruitment if they responded to the pregnancy baseline intrinsic motivation survey, did not decline future contact and data sharing, and had a confirmed live birth from their index PETALS pregnancy. PETALS-2 participants were enrolled at 6 months postpartum unless contact could not be made until 12 months postpartum. Exclusion criteria at the time of recruitment included: current or recent pregnancy (after the index PETALS pregnancy); demise of the index PETALS infant; current hospitalization; history of serious mental illness or eating disorder; and having moved or planning to move out of the geographic area. To adjust to research constraints following onset of the COVID-19 pandemic, the geographic eligibility criterion was dropped, and all assessments were administered remotely. All participants provided informed consent. The study was approved by the Kaiser Foundation Research Institute human subjects committee (institutional review board protocol 1295116).

### Data assessment

Participant characteristics including self-reported race and ethnicity, age, education, income, parity, and marital status were collected in the PETALS baseline self-report survey. Measured pre-pregnancy weight and height were sourced from electronic health records to calculate pre-pregnancy BMI. Using a longitudinal repeated measures design, PETALS-2 included in-person clinic visits and/or remote data collection assessments every six months at 6, 12, 18, and 24 months postpartum beginning in 2019. Weight to calculate postpartum BMI was assessed objectively via electronic health records (primary source), as well as study clinic visits, remote monitoring body weight scales (BodyTrace), or survey self-report, as previously described (Supplementary Materials).^15,17^

#### Intrinsic Motivation

Intrinsic motivation was assessed separately for healthy eating, physical activity, and self-weighing during pregnancy and at all postpartum time points.^15^ Intrinsic motivation for healthy eating and self-weighing were assessed with the 6-item Attitudes Toward Healthy Foods and 6-item Attitudes Toward Weighing surveys, respectively.^13^ Intrinsic motivation for physical activity was assessed with the validated 4-item Intrinsic Regulation subscale of the Behavioral Regulation in Exercise Questionnaire-Version 2 (BREQ-2).^18^ All intrinsic motivation items were on a 5-point Likert scale, with higher mean scores indicative of greater motivation. We also calculated a single composite intrinsic motivation score by averaging the intrinsic motivation scores for all three health behaviors, as described previously.^15^

#### Self-efficacy

We examined whether potential associations between intrinsic motivation and study outcomes remained after accounting for self-efficacy, a common focus of existing behavioral interventions. Drawing from social cognitive theory,^19^ self-efficacy represents an individual’s belief in their personal capability to engage in behaviors despite barriers.^20^ As a widely targeted driver of health behavior, we assessed self-efficacy separately for healthy eating (3 items), physical activity (2 items), and weight control (3 items) using behavior-specific subscales of a validated measure developed by Hofstetter et al. and adapted by Kendall et al.^21,22^ Administered during pregnancy and at all postpartum time points, higher mean scores on a 5-point Likert scale for each subscale indicate greater self-efficacy.

#### Health Behavior Outcomes

Healthy eating, physical activity, and self-weighing behaviors were assessed individually using validated self-report measures.

Diet was assessed using a modified version of the Block Food Frequency Questionnaire (FFQ) at all postpartum time points except 18 months (due to its relatively high participant burden).^23^ The FFQ was modified previoiusly to assess prenatal supplements^17^ and items consumed by the ethnically diverse target population.^24^ FFQ data were used to quantify diet quality with the Healthy Eating Index-2015 (HEI-2015), the most recent HEI at the time of data collection.^25^ On a 100-point scale, the thirteen diet components of the HEI-2015 are based on the 2015-2020 USDA Dietary Guidelines for Americans; HEI-2015 scores ≥70 or ≥80 are indicative of good to excellent diet quality depending on the population. We considered healthy eating as meeting or exceeding the PETALS-2 sample-specific overall 75^th^ percentile (*P75*) of total HEI-2015 score, given the lack of an appropriate postpartum reference population for dietary data comparisons. While FFQ data are not generally intended to calculate daily energy intake, HEI analyses excluded FFQ observations corresponding to implausible energy intake (<400 or >6,000 kcal/day), as previously described.^17^

Physical activity was assessed using two validated measures, to ascertain a) attainment of guideline-recommended moderate-to-vigorous physical activity (MVPA), and b) volume of MVPA and sedentary behavior. First, attainment of at least 150 minutes/week of leisure-time MVPA was assessed using the single-item Stanford Leisure-Time Activity Categorical Item (L-Cat) at all postpartum time points.^2,26–28^ Respondents select one of six descriptive options corresponding to weekly duration and intensity of leisure-time physical activity, which map onto the pre-specified threshold of national guidelines. Second, weekly time engaged in MVPA and sedentary behavior were estimated using the 35-item Pregnancy Physical Activity Questionnaire (PPAQ), which has been used in the postpartum period.^29,30^ Respondents report time spent in population-appropriate activities in the previous two months, with each activity corresponding to a specific activity level (sedentary, light, moderate, vigorous). The instrument was slightly modified to reflect newer technologies (e.g., “watching TV or a video” was updated to “watching TV, a movie, or video clip^”^). The PPAQ was administered at all postpartum time points except for 18 months due to its relatively high participant burden. MVPA was expressed in total minutes/week, having doubled vigorous-intensity physical activity time to normalize activity by intensity.^1,26,31^ Sedentary behavior was expressed as MET-hours/week, calculated by multiplying weekly sedentary activity time by respective Compendium-based MET values (ranging from 1-1.5 METs).^32^ Both MVPA and sedentary behavior were categorized as meeting or exceeding the PETALS-2 sample-specific overall *P75*.

Self-weighing, a standard weight management technique, was assessed at all postpartum time points with a single question adapted from the EARLY Self-Weighing Questionnaire and the Stability First trial.^13,30^ Respondents select their frequency of self-weighing in the past month, ranging from “never” to “several times a day.” We pre-specified a threshold for regular self-weighing as at least once per week.

#### Life’s Essential 8 Outcome

As a comprehensive assessment of cardiovascular health, we examined LE8 scores at 6, 12, 18, and 24 months postpartum. LE8 is comprised of eight components: diet, physical activity, sleep, nicotine use, BMI, cholesterol, glucose, and blood pressure. Components were assessed via self-report or objective measurements and scored individually (Supplementary Materials). In concordance with LE8 scoring recommendations^1^ and prior research calculating LE8 scores based on available data,^33^ LE8 components at each timepoint were averaged to calculate an LE8 score ranging from 0 to 100, analyzed as a continuous variable. We set a minimum data availability threshold of at least four available components to calculate an LE8 score. Higher LE8 scores represent better cardiovascular health (i.e., lower cardiovascular risk), with LE8 80-100 indicating optimal, 50-79 moderate, and 0-49 poor cardiovascular health.^1^

### Statistical Analyses

We used modified Poisson regression models with robust standard errors to estimate adjusted risk ratios (RR) and 95% confidence intervals (CI) for longitudinal associations between each behavior-specific measure of intrinsic motivation and pre-specified thresholds for corresponding healthy eating, physical activity, and self-weighing behaviors. Longitudinal analyses retained the temporal sequence of intrinsic motivation associated with non-concurrent health behavior by lagging intrinsic motivation to the immediate subsequent time point. Models included postpartum time point and the interaction of intrinsic motivation-by-time point to estimate time-specific associations. Model covariates included maternal age at delivery, pre-pregnancy BMI, race/ethnicity, education, and parity (model 1). To examine whether intrinsic motivation is associated with subsequent health behavior beyond the effects of self-efficacy, an additional set of models further adjusted for each behavior-specific self-efficacy score (model 2).

We fit a linear regression model using generalized estimating equations to examine the longitudinal association between the composite measure of intrinsic motivation and subsequent postpartum LE8 scores. As described above, analysis retained the temporal sequence of intrinsic motivation associated with non-concurrent LE8 scores by lagging intrinsic motivation to the immediate subsequent time point. The model used postpartum time point and the interaction of intrinsic motivation-by-time point to estimate time-specific mean differences. Model covariates included maternal age at delivery, pre-pregnancy BMI, race/ethnicity, education, and parity.

Participants who experienced an intervening pregnancy during the study period contributed data until pregnancy onset.^15^ Analyses were conducted using SAS version 9.4 (SAS Institute). B.P.S. had full access to all the data in the study and takes responsibility for its integrity and the data analysis.

## Results

A total of 472 individuals responded to the baseline PETALS assessment of intrinsic motivation in 2017-2018. Of those, 450 were potentially eligible for PETALS-2 and received a recruitment contact; 32 did not meet eligibility criteria and 107 declined participation or could not be reached. A total of 311 enrolled, yielding a recruitment rate of 74%. Participant characteristics, summarized in Table 1, show that 91% were aged 25-44 years, 70% self-identified with a racial or ethnic minority background, 36% did not have a college degree, and 45% had a pre-pregnancy BMI of 18.5-24.9 kg/m^2^.

**Table 1.** Characteristics of participants in the Pregnancy, Environment, and Lifestyle Study-2 (PETALS-2; *N*=311)

| **Participant Characteristics** | **n (%)** |
| --- | --- |
| Age at delivery (years) |  |
| 18-24 | 27 (8.7) |
| 25-29 | 70 (22.5) |
| 30-34 | 125 (40.2) |
| 35-39 | 72 (23.2) |
| 40-44 | 17 (5.5) |
| Race or ethnicity |  |
| Asian/Pacific Islander | 67 (21.5) |
| Black/African American | 22 (7.1) |
| Hispanic | 84 (27.0) |
| More than one race/ethnicity | 44 (14.1) |
| Non-Hispanic White | 94 (30.2) |
| Education |  |
| High school or less | 34 (10.9) |
| Some college | 78 (25.1) |
| College graduate | 101 (32.5) |
| Post-graduate | 98 (31.5) |
| Marital status |  |
| Married/civil union | 228 (73.3) |
| Not married; living with a partner | 43 (13.8) |
| Divorced/separated | 3 (1.0) |
| Single | 37 (11.9) |
| Household income (\$) | |
| < 50,000 | 63 (20.5) |
| 50,000-99,000 | 89 (28.9) |
| 100,000-149,999 | 60 (19.5) |
| ≥ 150,000 | 96 (31.2) |
| Unknown | 3 (1.0) |
| Parity |  |
| 0 | 160 (51.5) |
| 1 | 106 (34.1) |
| ≥ 2 | 41 (13.2) |
| Pre-pregnancy BMI, kg/m<sup>2</sup> |  |
| < 18.5 | 4 (1.3) |
| 18.5-24.9 | 140 (45.0) |
| 25.0-29.9 | 93 (29.9) |
| ≥ 30.0 | 74 (23.8) |

*Abbreviations:* BMI, body mass index. Missing race/ethnicity, education, marital status, and parity data for $n=4$ participants. Missing household income for $n=7$ participants.

Intrinsic motivation descriptive data are presented in Table 2. As previously reported, internal consistency reliability was adequate for each intrinsic motivation scale.^15^ Self-efficacy subscales also demonstrated adequate internal consistency in the analytic sample (Supplementary Materials). Intrinsic motivation was correlated with self-efficacy across time points, suggesting they are related but non-overlapping constructs (Supplementary Materials).

**Table 2.** Intrinsic motivation behavior-specific and composite scores by time point in the Pregnancy, Environment, and Lifestyle Study-2 (PETALS-2)

|  | Baseline |  | Postpartum time point |  |  |  |  |  |  |  |
| --- | --- | --- | --- | --- | --- | --- | --- | --- | --- | --- |
|  | Pregnancy |  | 6 months |  | 12 months |  | 18 months |  | 24 months |  |
|  | *n* | Mean (SD) | *n* | Mean (SD) | *n* | Mean (SD) | *n* | Mean (SD) | *n* | Mean (SD) |
| Intrinsic motivation for healthy eating | 305 | 3.87 (0.7) | 151 | 3.89 (0.7) | 141 | 3.83 (0.8) | 146 | 3.77 (0.8) | 154 | 3.77 (0.8) |
| Intrinsic motivation for physical activity | 306 | 3.32 (1.1) | 151 | 3.44 (1.0) | 140 | 3.38 (1.0) | 146 | 3.24 (1.1) | 154 | 3.38 (1.1) |
| Intrinsic motivation for self-weighing | 306 | 2.47 (0.9) | 151 | 2.48 (0.8) | 141 | 2.47 (0.9) | 146 | 2.48 (0.9) | 154 | 2.60 (0.9) |
| Intrinsic motivation composite | 306 | 3.22 (0.6) | 151 | 3.27 (0.6) | 141 | 3.23 (0.6) | 146 | 3.16 (0.7) | 154 | 3.25 (0.7) |

### Healthy Eating

Mean postpartum HEI-2015 scores peaked at 12 months postpartum (71.3; SD=10.2) compared to a low at 24 months (70.1; SD=9.5), with an overall P*75* of 75.2 across all time points (Table 3). Table 4 displays regression results of intrinsic motivation for healthy eating and HEI-2015 scores by time point. In model 1, adjusting for sociodemographic and clinical factors, each one-unit higher intrinsic motivation score for healthy eating was associated with a greater likelihood of subsequently reaching the HEI sample-specific *P75* at 6 months (RR=1.35; 95% CI=1.02, 1.78) and 24 months postpartum (1.46; 1.12, 1.90). In model 2, when adjusting for self-efficacy, the association was attenuated at 6 months and remained at 24 months postpartum (1.42; 1.07, 1.88).

**Table 3.**
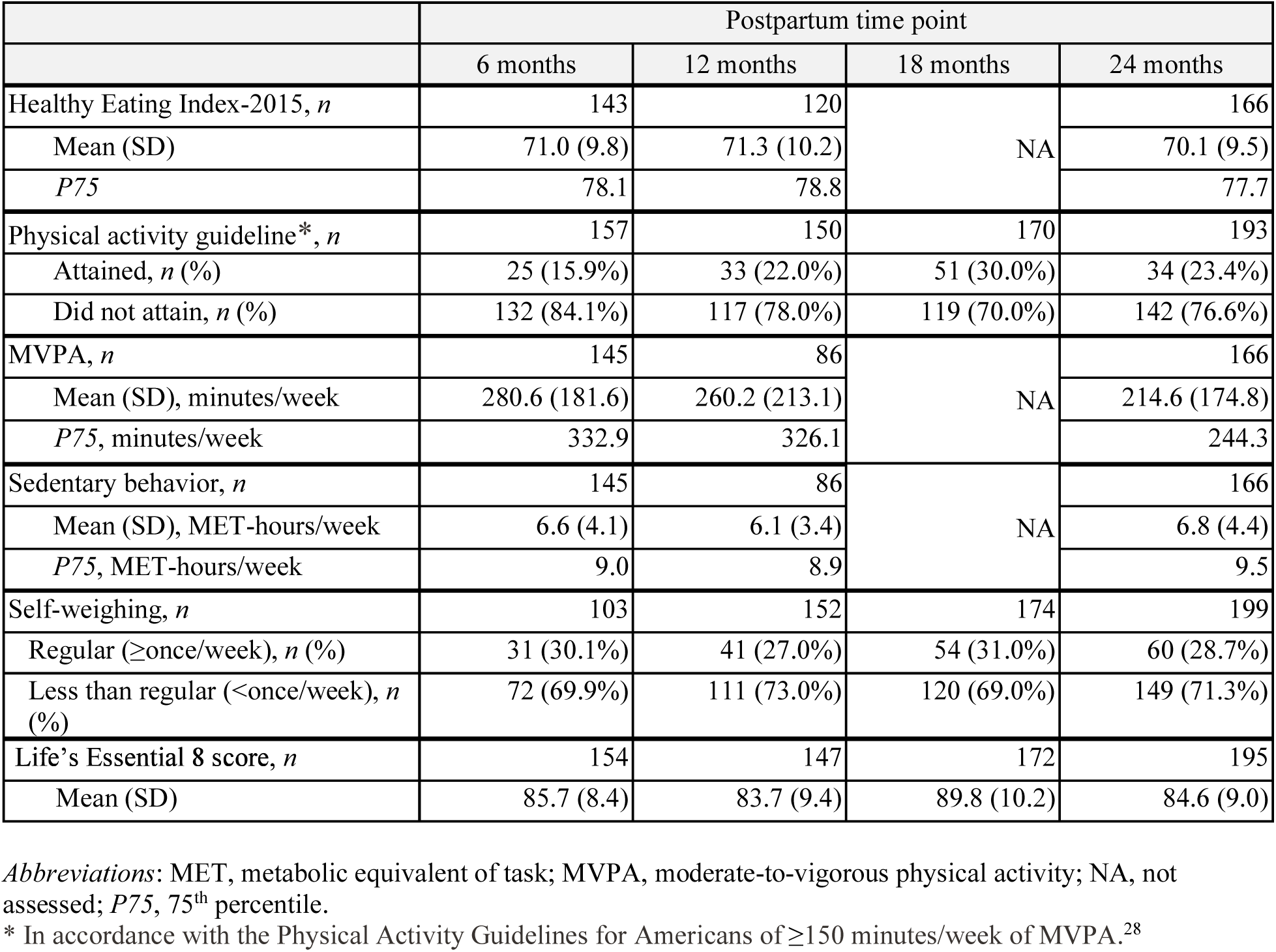
Health behavior and Life’s Essential 8 outcome scores by postpartum time point in the Pregnancy, Environment, and Lifestyle Study-2 (PETALS-2)

|  | Postpartum time point |  |  |  |
| --- | --- | --- | --- | --- |
|  | 6 months | 12 months | 18 months | 24 months |
| Healthy Eating Index-2015, *n* | 143 | 120 | NA | 166 |
| Mean (SD) | 71.0 (9.8) | 71.3 (10.2) |  | 70.1 (9.5) |
| *P75* | 78.1 | 78.8 |  | 77.7 |
| Physical activity guideline*, *n* | 157 | 150 | 170 | 193 |
| Attained, *n* (%) | 25 (15.9%) | 33 (22.0%) | 51 (30.0%) | 34 (23.4%) |
| Did not attain, *n* (%) | 132 (84.1%) | 117 (78.0%) | 119 (70.0%) | 142 (76.6%) |
| MVPA, *n* | 145 | 86 | NA | 166 |
| Mean (SD), minutes/week | 280.6 (181.6) | 260.2 (213.1) |  | 214.6 (174.8) |
| *P75*, minutes/week | 332.9 | 326.1 |  | 244.3 |
| Sedentary behavior, *n* | 145 | 86 | NA | 166 |
| Mean (SD), MET-hours/week | 6.6 (4.1) | 6.1 (3.4) |  | 6.8 (4.4) |
| *P75*, MET-hours/week | 9.0 | 8.9 |  | 9.5 |
| Self-weighing, *n* | 103 | 152 | 174 | 199 |
| Regular (≥once/week), *n* (%) | 31 (30.1%) | 41 (27.0%) | 54 (31.0%) | 60 (28.7%) |
| Less than regular ($<$once/week), *n* (%) | 72 (69.9%) | 111 (73.0%) | 120 (69.0%) | 149 (71.3%) |
| Life's Essential 8 score, *n* | 154 | 147 | 172 | 195 |
| Mean (SD) | 85.7 (8.4) | 83.7 (9.4) | 89.8 (10.2) | 84.6 (9.0) |

*Abbreviations:* MET, metabolic equivalent of task; MVPA, moderate-to-vigorous physical activity; NA, not assessed; *P75*, 75<sup>th</sup> percentile.
\* In accordance with the Physical Activity Guidelines for Americans of $\geq 150$ minutes/week of MVPA.<sup>28</sup>

**Table 4.**
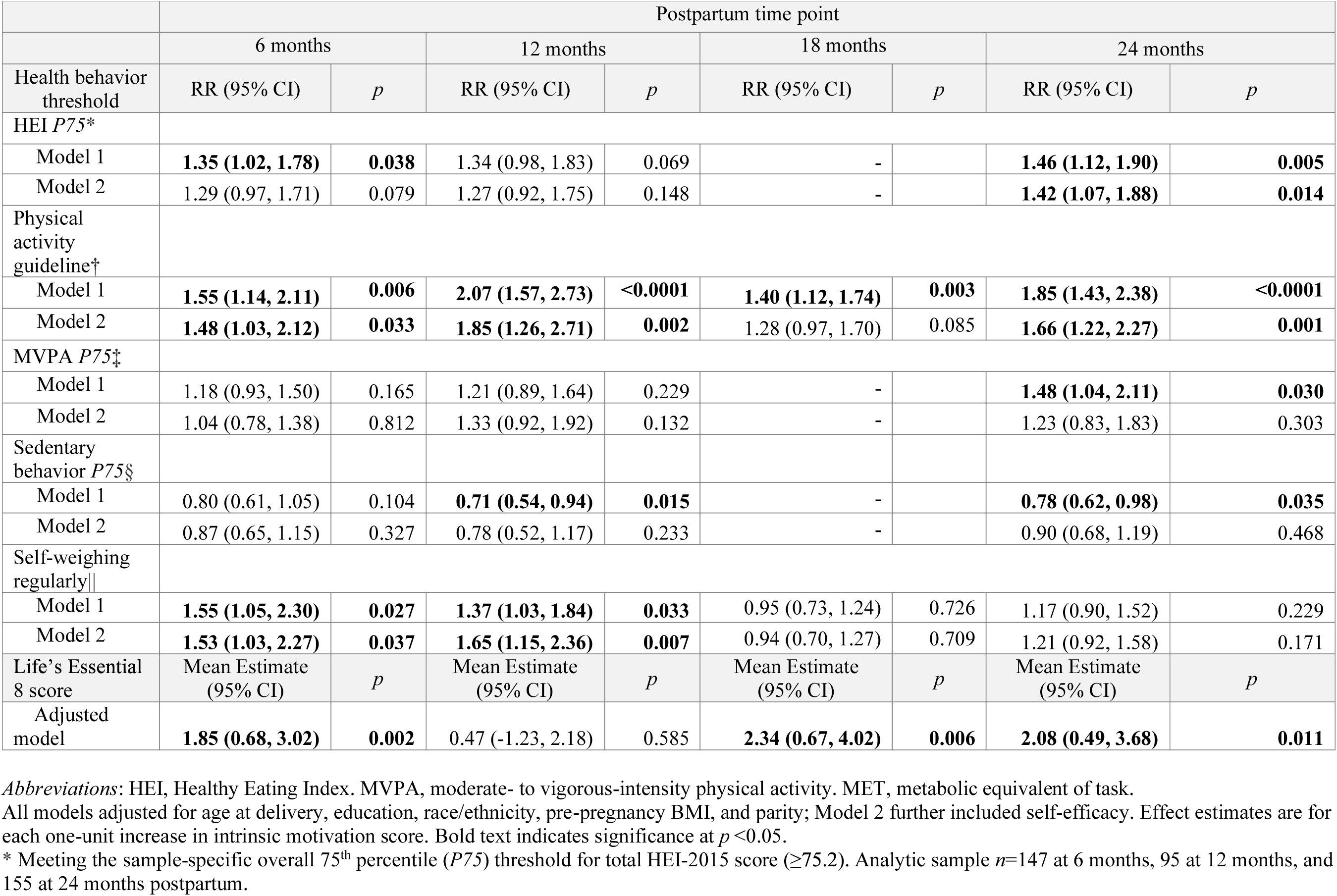
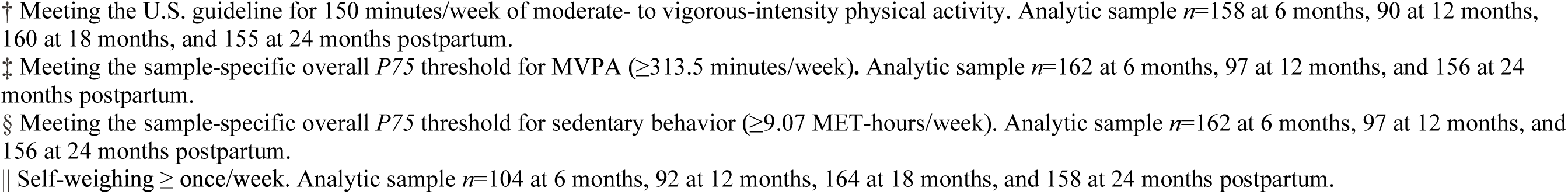
Longitudinal associations between intrinsic motivation for healthy eating, physical activity, and self-weighing and corresponding health behavior thresholds and continuous Life’s Essential 8 scores, by postpartum time point.

|  | Postpartum time point |  |  |  |  |  |  |  |
| --- | --- | --- | --- | --- | --- | --- | --- | --- |
|  | 6 months |  | 12 months |  | 18 months |  | 24 months |  |
| Health behavior threshold | RR (95% CI) | *p* | RR (95% CI) | *p* | RR (95% CI) | *p* | RR (95% CI) | *p* |
| HEI *P75* * |  |  |  |  |  |  |  |  |
| Model 1 | **1.35 (1.02, 1.78)** | **0.038** | 1.34 (0.98, 1.83) | 0.069 | - |  | **1.46 (1.12, 1.90)** | **0.005** |
| Model 2 | 1.29 (0.97, 1.71) | 0.079 | 1.27 (0.92, 1.75) | 0.148 | - |  | **1.42 (1.07, 1.88)** | **0.014** |
| Physical activity guideline† |  |  |  |  |  |  |  |  |
| Model 1 | **1.55 (1.14, 2.11)** | **0.006** | **2.07 (1.57, 2.73)** | **<0.0001** | **1.40 (1.12, 1.74)** | **0.003** | **1.85 (1.43, 2.38)** | **<0.0001** |
| Model 2 | **1.48 (1.03, 2.12)** | **0.033** | **1.85 (1.26, 2.71)** | **0.002** | 1.28 (0.97, 1.70) | 0.085 | **1.66 (1.22, 2.27)** | **0.001** |
| MVPA *P75*‡ |  |  |  |  |  |  |  |  |
| Model 1 | 1.18 (0.93, 1.50) | 0.165 | 1.21 (0.89, 1.64) | 0.229 | - |  | **1.48 (1.04, 2.11)** | **0.030** |
| Model 2 | 1.04 (0.78, 1.38) | 0.812 | 1.33 (0.92, 1.92) | 0.132 | - |  | 1.23 (0.83, 1.83) | 0.303 |
| Sedentary behavior *P75*§ |  |  |  |  |  |  |  |  |
| Model 1 | 0.80 (0.61, 1.05) | 0.104 | **0.71 (0.54, 0.94)** | **0.015** | - |  | **0.78 (0.62, 0.98)** | **0.035** |
| Model 2 | 0.87 (0.65, 1.15) | 0.327 | 0.78 (0.52, 1.17) | 0.233 | - |  | 0.90 (0.68, 1.19) | 0.468 |
| Self-weighting regularly\|\| |  |  |  |  |  |  |  |  |
| Model 1 | **1.55 (1.05, 2.30)** | **0.027** | **1.37 (1.03, 1.84)** | **0.033** | 0.95 (0.73, 1.24) | 0.726 | 1.17 (0.90, 1.52) | 0.229 |
| Model 2 | **1.53 (1.03, 2.27)** | **0.037** | **1.65 (1.15, 2.36)** | **0.007** | 0.94 (0.70, 1.27) | 0.709 | 1.21 (0.92, 1.58) | 0.171 |
| Life's Essential 8 score | Mean Estimate (95% CI) | *p* | Mean Estimate (95% CI) | *p* | Mean Estimate (95% CI) | *p* | Mean Estimate (95% CI) | *p* |
| Adjusted model | **1.85 (0.68, 3.02)** | **0.002** | 0.47 (-1.23, 2.18) | 0.585 | **2.34 (0.67, 4.02)** | **0.006** | **2.08 (0.49, 3.68)** | **0.011** |

*Abbreviations:* HEI, Healthy Eating Index. MVPA, moderate- to vigorous-intensity physical activity. MET, metabolic equivalent of task.
All models adjusted for age at delivery, education, race/ethnicity, pre-pregnancy BMI, and parity; Model 2 further included self-efficacy. Effect estimates are for each one-unit increase in intrinsic motivation score. Bold text indicates significance at $p < 0.05$ .
\* Meeting the sample-specific overall 75<sup>th</sup> percentile (*P75*) threshold for total HEI-2015 score ( $\geq 75.2$ ). Analytic sample $n=147$ at 6 months, 95 at 12 months, and 155 at 24 months postpartum.
- † Meeting the U.S. guideline for 150 minutes/week of moderate- to vigorous-intensity physical activity. Analytic sample $n=158$ at 6 months, 90 at 12 months, 160 at 18 months, and 155 at 24 months postpartum. - ‡ Meeting the sample-specific overall $P75$ threshold for MVPA ( $\geq 313.5$ minutes/week). Analytic sample $n=162$ at 6 months, 97 at 12 months, and 156 at 24 months postpartum. - § Meeting the sample-specific overall $P75$ threshold for sedentary behavior ( $\geq 9.07$ MET-hours/week). Analytic sample $n=162$ at 6 months, 97 at 12 months, and 156 at 24 months postpartum. - || Self-weighting ≥ once/week. Analytic sample $n=104$ at 6 months, 92 at 12 months, 164 at 18 months, and 158 at 24 months postpartum.

### Physical Activity

The proportion of participants who attained physical activity guidelines ranged from 15.9% at 6 months postpartum to 30.0% at 18 months postpartum (Table 3). In model 1, each one-unit higher intrinsic motivation score for physical activity was associated with a greater likelihood of subsequently attaining the physical activity guideline at 6 months (RR=1.55; 95% CI=1.14, 2.11), 12 months (2.07; 1.57, 2.73), 18 months (1.40; 1.12, 1.74), and 24 months postpartum (1.85; 1.43, 2.38; Table 4). In model 2, adjusting for self-efficacy, the association remained at 6 months (1.48; 1.03, 2.12), 12 months (1.85; 1.26, 2.71), and 24 months postpartum (1.66; 1.22, 2.27), but was attenuated at 18 months.

Mean postpartum MVPA peaked at 6 months postpartum (280.8 minutes/week; SD=181.6) compared to a low at 24 months postpartum (214.6 minutes/week; SD=174.8), with an overall *P75* of 313.5 minutes/week (Table 3). In model 1, each one-unit higher intrinsic motivation score for physical activity was associated with a greater likelihood of subsequently reaching the MVPA sample-specific *P75* at 24 months postpartum (1.48; 1.04, 2.11), but not after adjusting for self-efficacy in model 2 (Table 4).

Mean postpartum sedentary behavior peaked at 24 months postpartum (6.8 MET-hours/week; SD=4.4) compared to a low at 12 months postpartum (6.1 MET-hours/week; SD=3.4) at 12 months postpartum, with an overall *P75* of 9.5 MET-hours/week (Table 3). In model 1, each one-unit higher intrinsic motivation score for physical activity was associated with a lower likelihood of subsequently reaching the sample *P75* for sedentary behavior at 12 months (0.71; 0.54, 0.94) and 24 months postpartum (0.78; 0.62, 0.98; Table 4), but not after adjusting for self-efficacy in model 2.

### Self-Weighing

The proportion of participants who endorsed regular postpartum self-weighing (at least once weekly) ranged from 27.0% at 12 months to 31.0% at 18 months postpartum (Table 3). In model 1, each one-unit higher intrinsic motivation score for self-weighing was associated with a greater likelihood of subsequent regular self-weighing at 6 months (1.55; 1.05, 2.30) and 12 months postpartum (1.37; 1.03, 1.84; Table 4). In model 2, after adjusting for self-efficacy, this association remained at 6 months (1.53; 1.03, 2.27) and 12 months postpartum (1.65; 1.15, 2.36).

### Life’s Essential 8

Mean postpartum LE8 scores ranged from 83.7 (SD=9.4) at 12 months postpartum to 89.8 (SD=10.2) at 18 months postpartum (Table 3). Descriptive data on the Life’s Essential 8 (LE8) component scores used to calculate overall LE8 scores are presented in Supplementary Materials. In the adjusted model (Table 4), each one-unit higher composite intrinsic motivation score was associated with higher subsequent LE8 scores at 6 months (mean estimate=1.85; 95% CI=0.68, 3.02), 18 months (2.34; 0.67, 4.02), and 24 months postpartum (2.08; 0.49, 3.68).

## Discussion

This study in a racially and ethnically diverse cohort indicates that higher levels of intrinsic motivation for healthy eating, physical activity, and self-weighing were longitudinally associated with subsequent postpartum engagement in each corresponding health behavior throughout the first two years postpartum. These behaviors include meeting 75^th^ percentiles for diet quality, MVPA, and sedentary behavior (where higher motivation was associated with lower likelihood of the latter), as well as attaining national physical activity guidelines and self-weighing regularly. Moreover, higher levels of a composite measure of intrinsic motivation, encompassing all three health behaviors, was longitudinally associated with higher postpartum LE8 scores, a comprehensive indicator of cardiovascular health. The postpartum period can influence lifelong health, as postpartum weight retention is a known risk for later cardiovascular disease.^4,5,34^ The present findings expand our understanding of factors that influence postpartum health behaviors and, in turn, postpartum cardiovascular health (Figure 1). These results may inform interventions aiming to improve lifelong cardiovascular health.

A novel finding of this research is that intrinsic motivation has a unique influence on health behaviors, even when accounting for self-efficacy. Evidence for this emerged for healthy eating, meeting national physical activity guidelines, and self-weighing regularly, at time points ranging from 6 to 24 months postpartum. Intrinsic motivation and self-efficacy are understood to be related, yet the relationship among intrinsic motivation, self-efficacy, and engagement in cardiovascular health behaviors in a postpartum population had not been described prior to this study. To ensure that our intrinsic motivation and self-efficacy assessments represented distinct constructs, we used validated measures, confirmed their internal consistency reliability (here and Brown et al.),^15^ and verified that intrinsic motivation and self-efficacy were related but not overlapping psychosocial constructs. The present findings suggest that intrinsic motivation is a unique contributor to health behaviors, one that could be integrated into interventions aimed at improving maternal cardiovascular health.

The longitudinal data collection, spanning from pregnancy to two years postpartum, provides insight into differentially timed associations through this period. Notably, after adjusting for self-efficacy, behavior-specific intrinsic motivation was associated with self-weighing in the first postpartum year, healthy eating at two years postpartum, and attaining physical activity guidelines throughout the first two years postpartum. These results could inform the delivery timing of intervention components aimed at promoting postpartum health behaviors. For example, intrinsic motivation may be particularly important for interventions focused on weight management and reducing postpartum weight retention within the first year postpartum.

Beyond individual health behaviors, the present analyses found that higher levels of a novel composite measure of intrinsic motivation were longitudinally associated with higher LE8 cardiovascular health scores over the first two years postpartum, specifically at 6, 18, and 24 months. While prior research has calculated perinatal LE8 scores, such cardiovascular health characterizations have been limited to pregnancy and the first six months after delivery.^31,35,36^ Of note, LE8 scores in our sample were higher (∼84-90 out of 100 possible points) than in other perinatal samples.^31,35,37^ This may reflect a healthier sample of participants. It may also have been due in part to our dichotomous scoring of blood lipids, blood glucose, and blood pressure based on self-reported diagnosis history, rather than continuous clinical measurements. Future characterizations of maternal cardiovascular health would benefit from objective clinical measurements of health factors.

The present study has several strengths that contribute to its interpretability. First, the lagged analysis structure enabled insight into longitudinal associations between earlier intrinsic motivation with subsequent health behaviors and cardiovascular health scores. The extended follow-up period and racially and ethnically diverse sample are additional strengths. Given the postpartum period is understudied, this study could support the development of interventions promoting postpartum health behaviors, especially among individuals from diverse racial and ethnic minority backgrounds who are disproportionately affected by postpartum weight retention.^38^ Finally, the analysis of LE8 scores contributes a novel characterization of cardiovascular health throughout the first two postpartum years of a woman’s life.

Several study limitations exist. First, data missingness arose given the long-term follow-up for over two years, particularly spanning the COVID-19 pandemic. Still, the repeated measures design and the statistical approaches to accommodate variation in the number and spacing of observations per participant, maximizing use of all available data, served to mitigate this limitation. Second, although racially and ethnically diverse, the sample was more homogenous in education, thus limiting generalizability. Third, self-report data collection methods (particularly for diet and physical activity) are subject to bias, including recall errors and social desirability bias.^39,40^ However, these were mitigated in part by using validated measures and percentile thresholds that ranked behavior engagement within the sample. Fourth, we used a sample-specific percentile threshold for LE8 diet scoring due to the lack of pregnancy and postpartum reference populations.^1^

Our results provide valuable insight into postpartum intrinsic motivation, health behaviors, and cardiovascular health in a diverse sample. Future research could examine the efficacy and real-world effectiveness of interventions to improve postpartum intrinsic motivation for cardiovascular health behaviors, using existing techniques^13^ or novel approaches. In the context of maternal cardiovascular health disparities, intentional sampling methods should be used when conducting these studies.^41^ Additionally, integrating psychological and behavioral strategies into multi-component interventions could strengthen short- and long-term impacts on maternal health.^42^

In summary, findings from the PETALS-2 cohort demonstrate that higher intrinsic motivation was longitudinally associated with greater engagement in healthy eating, physical activity, and self-weighing behaviors, and with a comprehensive assessment of cardiovascular health, across the extended postpartum period. These findings indicate that intrinsic motivation could be a psychological target for interventions designed to improve health behaviors in the years following childbirth. Further research is needed to develop and evaluate both the content and timing of postpartum interventions that could target intrinsic motivation, thereby improving our understanding of influences on cardiovascular health throughout the life course.

## Data Availability

Study data are available from the corresponding author upon reasonable request.

## Acknowledgments

The authors extend appreciation to the study participants for their contributions to this research, and to Siedah Garrison, MPH, CCRP for project administration. **Sources of Funding:** This work was supported by National Institutes of Health grant R01 HL142996 to S.D.B. and R01 ES019196 to A.F. Additional support came from grants K26 DK138246, P30 DK092924, the UPSTREAM Center, and residual class settlement funds in the matter of April Krueger v. Wyeth, Inc., Case No. 03-cv-2496 (US District Court, SD of Calif.). The authors collected, analyzed, and interpreted the data and drafted the manuscript independently from sponsors.

## Non-standard Abbreviations and Acronyms

AHA: American Heart Association
BREQ-2: Behavioral Regulation in Exercise Questionnaire-Version 2
HEI: Healthy Eating Index
L-Cat: Stanford Leisure-Time Activity Categorical Item
LE8: Life’s Essential 8
MVPA: Moderate-to-vigorous physical activity
PETALS-2: Pregnancy, Lifestyle and Environment Study-2
PPAQ: Pregnancy Physical Activity Questionnaire

